# Prognostic factors in spanish COVID-19 patients: A case series from barcelona

**DOI:** 10.1101/2020.06.18.20134510

**Authors:** Antoni Sisó-Almirall, Belchin Kostov, Minerva Mas-Heredia, Sergi Vilanova-Rotllan, Ethel Sequeira-Aymar, Mireia Sans-Corrales, Elisenda Sant-Arderiu, Laia Cayuelas-Redondo, Angela Martínez-Pérez, Noemí García Plana, August Anguita-Guimet, Jaume Benavent-Àreu

**Author notes:** Corresponding Author: Dr Antoni Sisó Almirall MD. PhD, President of the Catalan Society of Family and Community Medicine (CAMFiC)., Head of Research. CAP Les Corts, CAPSBE, Carrer Mejía Lequerica s/n. Barcelona 08028. Spain.

## Abstract

**Background:** In addition to the lack of COVID-19 diagnostic tests for the whole Spanish population, the current strategy is to identify the disease early to limit contagion in the community.

**Aim:** To determine clinical factors of a poor prognosis in patients with COVID-19 infection.

**Design and Setting:** Descriptive, observational, retrospective study in three primary healthcare centres with an assigned population of 100,000.

**Method:** Examination of the medical records of patients with COVID-19 infections confirmed by polymerase chain reaction.

**Results:** We included 322 patients (mean age 56.7 years, 50% female, 115 (35.7%) aged ≥ 65 years). The best predictors of ICU admission or death were greater age, male sex (OR=2.99; 95%CI=1.55 to 6.01), fever (OR=2.18; 95%CI=1.06 to 4.80), dyspnoea

(OR=2.22; 95%CI=1.14 to 4.24), low oxygen saturation (OR=2.94; 95%CI=1.34 to 6.42), auscultatory alterations (OR=2.21; 95%CI=1.00 to 5.29), heart disease (OR=4.37; 95%CI=1.68 to 11.13), autoimmune disease (OR=4.03; 95%CI=1.41 to 11.10), diabetes (OR=4.00; 95%CI=1.89 to 8.36), hypertension (OR=3.92; 95%CI=2.07 to 7.53), bilateral pulmonary infiltrates (OR=3.56; 95%CI=1.70 to 7.96), elevated lactate-dehydrogenase (OR=3.02; 95%CI=1.30 to 7.68), elevated C-reactive protein (OR=2.94; 95%CI=1.47 to 5.97), elevated D-dimer (OR=2.66; 95%CI=1.15 to 6.51) and low platelet count (OR=2.41; 95%CI=1.12 to 5.14). Myalgia or artralgia (OR=0.28; 95%CI=0.10 to 0.66), dysgeusia (OR=0.28; 95%CI=0.05 to 0.92) and anosmia (OR=0.23; 95%CI=0.04 to 0.75) were protective factors.

**Conclusion:** Determining the clinical, biological and radiological characteristics of patients with suspected COVID-19 infection will be key to early treatment and isolation and the tracing of contacts.

## INTRODUCTION

On 31 December 2019, the health authorities of Wuhan city (Hubei Province, China) reported a cluster of 27 cases of pneumonia of unknown aetiology with onset of symptoms on 8 December, including 7 severe cases, with a common exposure identified in a city market [1], which was closed on January 1, 2020. On 7 January 2020, the Chinese authorities identified a new *Coronaviridae* family virus, initially named coronavirus 2,019-nCoV and later coronavirus SARS-CoV-2 as the causal agent [2].

The genetic sequence was shared by the Chinese authorities on 12 January 2020. On January 19, the first case was detected in the USA, in Washington state [3]. On 30 January 2020, the World Health Organization declared the SARS-CoV-2 outbreak in China a public health emergency of international concern [4]. Subsequently, the outbreak has spread outside China, with Europe especially affected [5].

The first positive case diagnosed in Spain was confirmed on January 31, 2020 on the island of La Gomera, while the first death occurred on February 13 in Valencia city (the date was confirmed twenty days later). The first confirmed case in Barcelona was on 24 February, and from then until 20 April 2020, there have been 195,944 confirmed cases in Spain [6], the highest number in the European Union.

The most common signs of infection are respiratory symptoms: fever, cough and shortness of breath. In more severe cases, the infection may cause pneumonia, severe acute respiratory syndrome, renal failure and death [7]. Transmission appears to be mainly person-to-person via the airway through respiratory droplets measuring > 5 microns when the patient has respiratory symptoms (cough and sneezing) and contact with fomites [8]. Most estimates of the incubation period of COVID-19 range from 1 to 14 days, with most around five days. Evidence on the transmission of the virus before symptom onset is unclear. There is currently no specific treatment for COVID-19 infections. To date, the most important scientific efforts have focused on three areas: strategies to contain the spread of the disease, the initiation of clinical trials with antivirals and multiple therapies, and the design of a new vaccine, which is still unclear. These strategies include some of a community nature, where primary healthcare plays a central role in disease prevention and control [9]. Few studies have described the clinical characteristics of the disease, fewer the predictive factors, and virtually none have described the Mediterranean population compared with the rest of the world.

Therefore, this study aimed to describe the clinical, biological and radiological manifestations, the evolution, treatments and mortality rate of patients with COVID-19 infection in the population of Barcelona city and determine the most important predictors of a poor prognosis.

## MATERIALS AND METHODS

A multicentre, observational descriptive study was carried out in three urban primary healthcare centres serving an assigned population of 100,000, with one reference hospital. The first patients diagnosed with COVID-19 confirmed by polymerase chain reaction (PCR) from nasal and pharyngeal samples were included. Diagnostic confirmation was made in the hospital laboratories, as PCR is not available in primary healthcare centres. Signs and symptoms, the main available haematological and biochemical data and the results of imaging tests were recorded, as were comorbidities, the evolution, the hospitalization rate, intensive care unit (ICU) admission and the treatments received. Other variables recorded were the type of follow-up, the need for temporary work disability, and the source of possible contacts. The factors that determined a poor prognosis (hospitalization, ICU admission, death) were collected. The data were obtained from the electronic medical record. Missing data were collected by telephone interviews with patients when possible. Patients from nursing homes were excluded, as the rate of infections and mortality has been shown to be much higher than in the non-institutionalized population. The study was approved by the Ethics Committee of the Hospital Clinic of Barcelona (registration number HCB/2020/0525). The study was conducted according to the Helsinki Declaration and Spanish legislation on biomedical studies, data protection and respect for human rights.

### Statistical analysis

Categorical variables are presented as absolute frequencies and percentages (%) and continuous variables as means and standard deviations (SD). Predictors of death, ICU admission and hospitalization were determined using the student’s t test for continuous variables and Fisher’s exact test for categorical variables. The odds ratio was calculated for categorical predictors. Values of p<0.05 were considered statistically significant and 95% confidence intervals (CI) were calculated. The statistical analysis was performed using the R version 3.6.1. for Windows.

## RESULTS

### Clinical characteristics and comorbidities

We included 322 patients (mean age 56.7 years, 50% female, 115 (35.7%) aged ≥ 65 years). The mean time from symptom onset to the medical visit was 3.9 (SD 4.6) days. Clinical characteristics are shown in **Table 1**. Notably, 123 (38.2) were health workers (doctors, nurses, auxiliaries). The most frequent clinical symptoms were cough (73.9%), fever (63.8%), general malaise (43.5%), fatigue (30.7%), myalgia or arthralgia (30.1%), dyspnoea (25.5%), diarrhoea (23%), headache (20.8%), anosmia (17.4%) and dysgeusia (14.9%). Physical examination in 223 (69.3%) patients showed 69.1% had auscultatory alterations, 28.7% tachypnoea and 20.5% an oxygen saturation of ≤ 92%.

**Table 1.** Clinical and exploratory factors predicting hospitalization and ICU admission/death.

| Variables | Total<br>(n=322) | Death or ICU admission |  |  |  | Hospitalization |  |  |  |
| --- | --- | --- | --- | --- | --- | --- | --- | --- | --- |
|  |  | No (n=266) | Yes (n=56) | P | OR [95% CI] | No (n=164) | Yes (n=158) | P | OR [95% CI] |
| **Age — years** | 56.7 ± 17.8 | 54.3 ± 17.4 | 68.2 ± 14.9 | **<0.001** |  | 48.6 ± 16.0 | 65.1 ± 15.6 | **<0.001** |  |
| Distribution — no. (%) |  |  |  | **<0.001** |  |  |  | **<0.001** |  |
| 15–30 years | 32 (9.9) | 30 (11.3) | 2 (3.6) |  | Ref | 30 (18.3) | 2 (1.3) |  | Ref |
| 31–49 years | 92 (28.6) | 88 (33.1) | 4 (7.1) |  | 0.68 [0.09-7.92] | 61 (37.2) | 31 (19.6) |  | 7.53 [1.72-69.31] |
| 50–64 years | 83 (25.8) | 71 (26.7) | 12 (21.4) |  | 2.52 [0.51-24.53] | 46 (28.0) | 37 (23.4) |  | 11.86 [2.71-109.05] |
| ≥65 years | 115 (35.7) | 77 (28.9) | 38 (67.9) |  | 7.33 [1.70-66.66] | 27 (16.5) | 88 (55.7) |  | 47.4 [10.9-434.7] |
| **Male — no. (%)** | 161 (50.0) | 121 (45.5) | 40 (71.4) | **0.001** | 2.99 [1.55-6.01] | 67 (40.9) | 94 (59.5) | **0.001** | 2.12 [1.33-3.40] |
| **Occupation — no. (%)** |  |  |  | **<0.001** |  |  |  | **<0.001** |  |
| Other type of exposure | 189 (58.7) | 133 (50.0) | 56 (100.0) |  | Ref | 48 (29.3) | 141 (89.2) |  | Ref |
| Health professional | 123 (38.2) | 123 (46.2) | 0 (0) |  | 0.00 [0.00-0.08] | 106 (64.6) | 17 (10.8) |  | 0.06 [0.03-0.10] |
| Other health workers | 10 (3.1) | 10 (3.8) | 0 (0) |  | 0.00 [0.00-1.11] | 10 (6.1) | 0 (0) |  | 0.00 [0.00-0.16] |
| **Smoking (ex-smoker/smoker) — no./total no. (%)** | 81/260 (31.2) | 60/212 (28.3) | 21/48 (43.8) | 0.057 | 1.96 [0.98-3.93] | 31/119 (26.1) | 50/141 (35.5) | 0.109 | 1.56 [0.88-2.77] |
| **Temperature at admission — °C†** | 37.6 ± 0.8 | 37.6 ± 0.8 | 37.8 ± 0.7 | **0.030** |  | 37.3 ± 0.8 | 37.9 ± 0.7 | **<0.001** |  |
|  | 194/304 |  |  |  |  | 73/152 | 121/152 |  |  |
| Pacients with fever (≥37.5°C) — no./total no. (%) | (63.8) | 153/251 (61.0) | 41/53 (77.4) | **0.027** | 2.18 [1.06-4.80] | (48.0) | (79.6) | **<0.001** | 4.20 [2.47-7.27] |
| **Time from symptom onset to medical visit — days‡** | 3.9 ± 4.6 | 3.9 ± 4.8 | 3.9 ± 3.7 | 0.954 |  | 3.5 ± 4.9 | 4.3 ± 4.3 | 0.166 |  |
| **Symptoms — no. (%)¶** |  |  |  |  |  |  |  |  |  |
| Cough | 238 (73.9) | 198 (74.4) | 40 (71.4) | 0.620 | 0.86 [0.44-1.75] | 114 (69.5) | 124 (78.5) | 0.076 | 1.60 [0.94-2.74] |
| General malaise | 140 (43.5) | 114 (42.9) | 26 (46.4) | 0.658 | 1.16 [0.62-2.14] | 65 (39.6) | 75 (47.5) | 0.178 | 1.37 [0.86-2.19] |
| Fatigue | 99 (30.7) | 82 (30.8) | 17 (30.4) | 1.000 | 0.98 [0.49-1.89] | 41 (25.0) | 58 (36.7) | **0.029** | 1.74 [1.05-2.90] |
| Myalgia or arthralgia | 97 (30.1) | 90 (33.8) | 7 (12.5) | **0.001** | 0.28 [0.10-0.66] | 53 (32.3) | 44 (27.8) | 0.397 | 0.81 [0.49-1.34] |
| Dyspnoea | 82 (25.5) | 60 (22.6) | 22 (39.3) | **0.012** | 2.22 [1.14-4.24] | 24 (14.6) | 58 (36.7) | **<0.001** | 3.37 [1.91-6.08] |
| Diarrhoea | 74 (23.0) | 64 (24.1) | 10 (17.9) | 0.384 | 0.69 [0.29-1.48] | 29 (17.7) | 45 (28.5) | **0.024** | 1.85 [1.06-3.27] |
| Headache | 67 (20.8) | 62 (23.3) | 5 (8.9) | **0.018** | 0.32 [0.10-0.86] | 37 (22.6) | 30 (19.0) | 0.493 | 0.81 [0.45-1.43] |
| Anosmia | 56 (17.4) | 53 (19.9) | 3 (5.4) | **0.006** | 0.23 [0.04-0.75] | 45 (27.4) | 11 (7.0) | **<0.001** | 0.20 [0.09-0.41] |
| Dysgeusia | 48 (14.9) | 45 (16.9) | 3 (5.4) | **0.024** | 0.28 [0.05-0.92] | 35 (21.3) | 13 (8.2) | **0.001** | 0.33 [0.15-0.68] |
| Sore throat | 38 (11.8) | 33 (12.4) | 5 (8.9) | 0.648 | 0.69 [0.20-1.91] | 25 (15.2) | 13 (8.2) | 0.058 | 0.50 [0.23-1.06] |
| Blocked nose | 38 (11.8) | 34 (12.8) | 4 (7.1) | 0.360 | 0.53 [0.13-1.57] | 24 (14.6) | 14 (8.9) | 0.122 | 0.57 [0.26-1.20] |
| Nausea or vomiting | 38 (11.8) | 30 (11.3) | 8 (14.3) | 0.500 | 1.31 [0.49-3.16] | 15 (9.1) | 23 (14.6) | 0.167 | 1.69 [0.81-3.64] |
| Sputum production | 29 (9.0) | 21 (7.9) | 8 (14.3) | 0.130 | 1.94 [0.70-4.90] | 13 (7.9) | 16 (10.1) | 0.561 | 1.31 [0.57-3.07] |
| Chills | 21 (6.5) | 17 (6.4) | 4 (7.1) | 0.770 | 1.13 [0.26-3.65] | 4 (2.4) | 17 (10.8) | **0.003** | 4.80 [1.52-20.07] |
| Asthenia | 12 (3.7) | 10 (3.8) | 2 (3.6) | 1.000 | 0.95 [0.10-4.64] | 3 (1.8) | 9 (5.7) | 0.081 | 3.23 [0.79-18.90] |
| Chest pain | 9 (2.8) | 7 (2.6) | 2 (3.6) | 0.658 | 1.37 [0.14-7.46] | 7 (4.3) | 2 (1.3) | 0.174 | 0.29 [0.03-1.55] |
| **Alterations in physical examination — no./total no. (%)§** |  |  |  |  |  |  |  |  |  |
|  | 154/223 |  |  |  |  |  | 112/148 |  |  |
| Auscultatory alterations | (69.1) | 112/171 (65.5) | 42/52 (80.8) | **0.041** | 2.21 [1.00-5.29] | 42/75 (56.0) | (75.7) | **0.004** | 2.43 [1.29-4.60] |
| Tachypnoea | 64/223 (28.7) | 43/171 (25.1) | 21/52 (40.4) | **0.037** | 2.01 [1.00-4.05] | 15/75 (20.0) | 49/148 (33.1) | **0.043** | 1.97 [1.00-4.13] |
| Tachycardia | 29/223 (13.0) | 23/171 (13.5) | 6/52 (11.5) | 0.818 | 0.84 [0.26-2.29] | 7/75 (9.3) | 22/148 (14.9) | 0.296 | 1.69 [0.66-4.94] |
| Pharyngitis | 22/223 (9.9) | 22/171 (12.9) | 0/52 (0) | **0.003** | 0.00 [0.00-0.55] | 9/75 (12.0) | 13/148 (8.8) | 0.480 | 0.71 [0.26-1.98] |
| **Oxygen saturation ≤92%— no./total no. (%)** | 42/205 (20.5) | 24/154 (15.6) | 18/51 (35.3) | **0.005** | 2.94 [1.34-6.42] | 8/66 (12.1) | 34/139 (24.5) | **0.043** | 2.34 [1.00-6.25] |

Statistically significant differences in bold (p<0.05).
† Temperature distribution was: <37.5°C (36.2%), 37.5–38.0°C (18.4%), 38.1–39.0°C (38.8%) and >39.0°C (6.6%).
‡ In 10 (3.1%) patients data on period between symptom onset and medical visit were lacking.
¶ Symptoms with a frequency of < 5 patients were: disorientation (n=4), conjunctivitis (n=3), hemoptysis (n=2) and cutaneous lesions (n=2).
§ 223 (69.3%) patients had a physical examination. The alterations with a frequency of < 5 patients were: cutaneous lesions (n=2) and tonsillopharyngitis (n=1).
Ref: reference.

The best predictors of ICU admission and death were age, male sex (OR = 2.99; 95%CI = 1.55 to 6.01), dyspnoea (OR = 2.22; 95%CI = 1.14 to 4.24), fever (OR = 2.18; 95%CI = 1.06 to 4.80), auscultatory alterations (OR = 2.21; 95%CI = 1.00 to 5.29) and low oxygen saturation (OR = 2.94; 95%CI = 1.34 to 6.42). However, myalgia or arthralgia (OR = 0.28; 95%CI = 0.10 to 0.66), dysgeusia (OR = 0.28; 95%CI = 0.05 to 0.92) and anosmia (OR = 0.23; 95%CI = 0.04 to 0.75) were significant protective factors against ICU admission and death (**Fig 1**).

**Fig 1.**
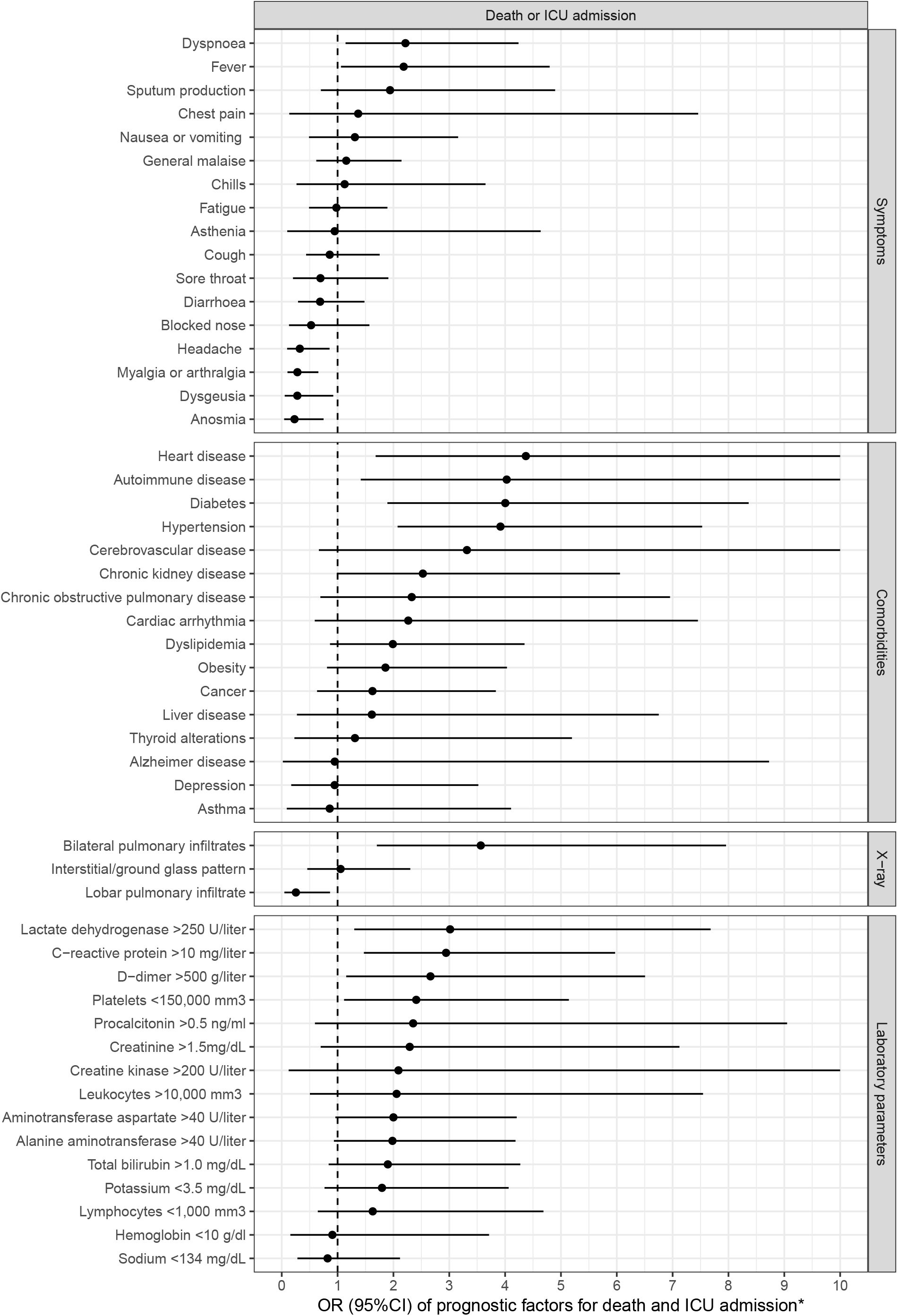
Prognostic factors for death and ICU admission. *The upper bounds of the confidence intervals were cut in 10.

Comorbidities were presented by 212 (65. 8%) patients: the most common were hypertension in 109 (33.9%), diabetes mellitus in 46 (14.3%), and obesity in 46 (14.3%) (**Table 2**). The most important predisposing factors for ICU admission and death were heart disease (OR = 4.37; 95%CI = 1.68 to 11.13), autoimmune disease (OR = 4.03; 95%CI = 1.41 to 11.10), diabetes (OR = 4.00; 95%CI = 1.89 to 8.36) and hypertension (OR = 3.92; 95%CI = 2.07 to 7.53) (**Fig 1**). Chills (OR = 4.80; 95%CI = 1.52 to 20.07), fever (OR = 4.20; 95%CI = 2.47 to 7.27), dyspnoea (OR = 3.37; 95%CI = 1.91 to 6.08), depression (OR = 9.08; 95%CI = 2.08 to 82.91), heart disease (OR = 6.10; 95%CI = 1.99 to 25.04) and chronic obstructive pulmonary disease (OR = 6.02; 95%CI = 1.67 to 32.89) are the best predictors of hospitalization (**Fig 2**).

**Table 2.** Comorbidities associated with hospitalization and ICU admission/death.

| Variables | Total<br>(n=322) | Death or ICU admission |  |  |  | Hospitalization |  |  |  |
| --- | --- | --- | --- | --- | --- | --- | --- | --- | --- |
|  |  | No (n=266) | Yes (n=56) | P | OR [95% CI] | No (n=164) | Yes (n=158) | P | OR [95% CI] |
| Comorbidities — no. (%)† |  |  |  |  |  |  |  |  |  |
| Any comorbidity | 212 (65.8) | 161 (60.5) | 51 (91.1) | <0.001 | 6.62 [2.54-21.96] | 82 (50.0) | 130 (82.3) | <0.001 | 4.62 [2.71-8.04] |
| Hypertension | 109 (33.9) | 75 (28.2) | 34 (60.7) | <0.001 | 3.92 [2.07-7.53] | 29 (17.7) | 80 (50.6) | <0.001 | 4.75 [2.80-8.24] |
| Diabetes | 46 (14.3) | 28 (10.5) | 18 (32.1) | <0.001 | 4.00 [1.89-8.36] | 13 (7.9) | 33 (20.9) | 0.001 | 3.06 [1.49-6.62] |
| Obesity | 46 (14.3) | 34 (12.8) | 12 (21.4) | 0.097 | 1.86 [0.81-4.03] | 13 (7.9) | 33 (20.9) | 0.001 | 3.06 [1.49-6.62] |
| Dyslipidemia | 44 (13.7) | 32 (12.0) | 12 (21.4) | 0.084 | 1.99 [0.86-4.35] | 18 (11.0) | 26 (16.5) | 0.194 | 1.60 [0.80-3.24] |
| Cancer | 37 (11.5) | 28 (10.5) | 9 (16.1) | 0.250 | 1.62 [0.63-3.83] | 13 (7.9) | 24 (15.2) | 0.054 | 2.08 [0.97-4.63] |
| Chronic kidney disease | 31 (9.6) | 21 (7.9) | 10 (17.9) | 0.042 | 2.53 [1.00-6.06] | 9 (5.5) | 22 (13.9) | 0.013 | 2.78 [1.18-7.10] |
| Heart disease | 25 (7.8) | 14 (5.3) | 11 (19.6) | 0.001 | 4.37 [1.68-11.13] | 4 (2.4) | 21 (13.3) | <0.001 | 6.10 [1.99-25.04] |
| Autoimmune disease | 21 (6.5) | 12 (4.5) | 9 (16.1) | 0.004 | 4.03 [1.41-11.10] | 7 (4.3) | 14 (8.9) | 0.116 | 2.18 [0.79-6.56] |
| Chronic obstructive pulmonary disease | 19 (5.9) | 13 (4.9) | 6 (10.7) | 0.114 | 2.33 [0.69-6.95] | 3 (1.8) | 16 (10.1) | 0.002 | 6.02 [1.67-32.89] |
| Depression | 18 (5.6) | 15 (5.6) | 3 (5.4) | 1.000 | 0.95 [0.17-3.52] | 2 (1.2) | 16 (10.1) | <0.001 | 9.08 [2.08-82.91] |
| Cardiac arrhythmia | 16 (5.0) | 11 (4.1) | 5 (8.9) | 0.168 | 2.27 [0.59-7.45] | 6 (3.7) | 10 (6.3) | 0.312 | 1.78 [0.57-6.10] |
| Thyroid alterations | 14 (4.3) | 11 (4.1) | 3 (5.4) | 0.717 | 1.31 [0.23-5.20] | 5 (3.0) | 9 (5.7) | 0.283 | 1.92 [0.56-7.45] |
| Asthma | 13 (4.0) | 11 (4.1) | 2 (3.6) | 1.000 | 0.86 [0.09-4.11] | 7 (4.3) | 6 (3.8) | 1.000 | 0.89 [0.24-3.16] |
| Liver disease | 12 (3.7) | 9 (3.4) | 3 (5.4) | 0.445 | 1.61 [0.27-6.75] | 5 (3.0) | 7 (4.4) | 0.568 | 1.47 [0.39-6.02] |
| Cerebrovascular disease | 10 (3.1) | 6 (2.3) | 4 (7.1) | 0.076 | 3.32 [0.66-14.56] | 2 (1.2) | 8 (5.1) | 0.057 | 4.30 [0.84-42.27] |
| Alzheimer disease | 6 (1.9) | 5 (1.9) | 1 (1.8) | 1.000 | 0.95 [0.02-8.73] | 1 (0.6) | 5 (3.2) | 0.115 | 5.30 [0.58-253.15] |

Statistically significant differences in bold (p<0.05).
† Comorbidities with a frequency of < 5 patients were: bronchiectasis (n=4), fibromyalgia (n=4), anemia (n=3), arthritis (n=3), HIV (n=2), syphilis (n=1) and tuberculosis (n=1).

**Fig 2.**
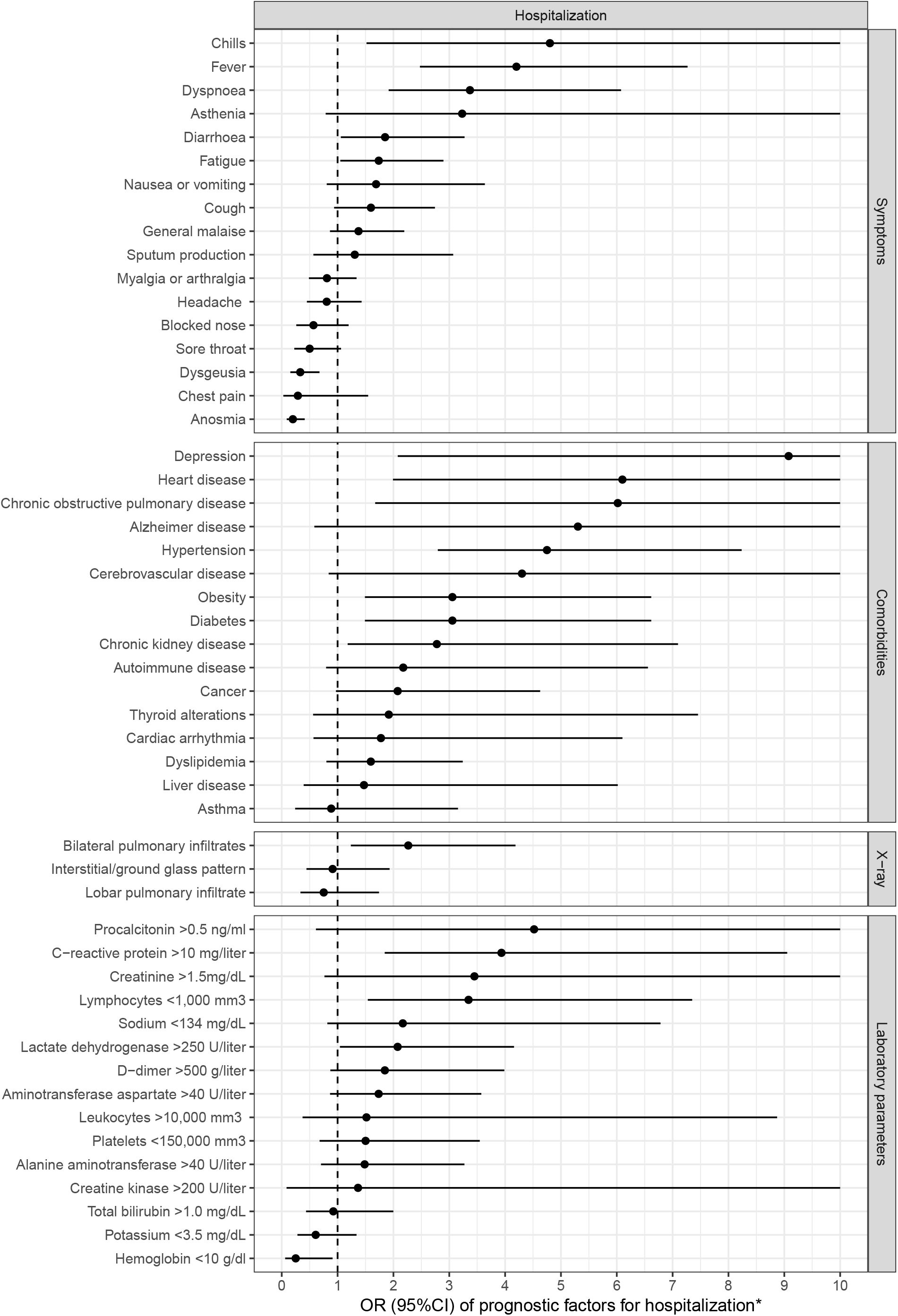
Prognostic factors for hospitalization. *The upper bounds of the confidence intervals were cut in 10.

### Imaging and laboratory tests

Chest X-ray was necessary in 227 patients (70.5%) and showed lobar pulmonary infiltrates in 35 (15.4%), bilateral pulmonary infiltrates in 129 (56.8%) and an interstitial pattern in 48 (21.1%) (**Table 3**). Chest CT was required in 28 patients and pulmonary ultrasound in 10 (3.1%). Biologically, 171 (81.4%) of 210 patients had lymphopenia (< 1,000 mm3). Likewise, 60.8% had a lactate dehydrogenase (LDH) > 250 U/ml and liver test alterations were common: elevated AST/GOT in 41.4% and ALT/GPT in 32.4%. In 86 (52.1%) of 165 cases D-dimer was elevated (> 500g/L). The most important factors of a poor prognosis were bilateral pulmonary infiltrates (OR = 3.56; 95%CI = 1.70 to 7.96), elevated lactate-dehydrogenase (OR = 3.02; 95%CI = 1.30 to 7.68), elevated C-reactive protein (OR = 2.94; 95%CI = 1.47 to 5.97), elevated D-dimer (OR = 2.66; 95%CI = 1.15 to 6.51) and low platelet count (OR = 2.41; 95%CI = 1.12 to 5.14) (**Fig 1**).

**Table 3.** Analytical and radiological predictors of hospitalization and ICU admission/death.

| Variables | Total<br>(n=322) | Death or ICU admission (n=56) |  |  |  | Hospitalization (n=158) |  |  |  |
| --- | --- | --- | --- | --- | --- | --- | --- | --- | --- |
|  |  | No (n=266) | Yes (n=56) | P | OR [95% CI] | No (n=164) | Yes (n=158) | P | OR [95% CI] |
| **Alterations in chest X-ray — no./total no. (%)†** |  |  |  |  |  |  |  |  |  |
| Bilateral pulmonary infiltrates | 129/227 (56.8) | 86/172 (50.0) | 43/55 (78.2) | **<0.001** | 3.56 [1.70-7.96] | 31/72 (43.1) | 98/155 (63.2) | **0.006** | 2.27 [1.24-4.19] |
| Interstitial/ground glass pattern | 48/227 (21.1) | 36/172 (20.9) | 12/55 (21.8) | 0.852 | 1.05 [0.46-2.30] | 16/72 (22.2) | 32/155 (20.6) | 0.862 | 0.91 [0.44-1.93] |
| Lobar pulmonary infiltrate | 35/227 (15.4) | 32/172 (18.6) | 3/55 (5.5) | **0.018** | 0.25 [0.05-0.86] | 13/72 (18.1) | 22/155 (14.2) | 0.438 | 0.75 [0.34-1.74] |
| **Alterations in chest CAT scan — no./total no. (%)‡** |  |  |  |  |  |  |  |  |  |
| Bilateral pulmonary infiltrates | 14/28 (50.0) | 6/17 (35.3) | 8/11 (72.7) | 0.120 | 4.60 [0.74-37.7] | 3/5 (60.0) | 11/23 (47.8) | 1.000 | 0.62 [0.04-6.54] |
| Interstitial/ground glass pattern | 13/28 (46.4) | 10/17 (58.8) | 3/11 (27.3) | 0.137 | 0.28 [0.03-1.70] | 2/5 (40.0) | 11/23 (47.8) | 1.000 | 1.36 [0.13-19.19] |
| **Laboratory parameters — no./total no. (%)** |  |  |  |  |  |  |  |  |  |
| Leukocytes >10,000 mm3 | 13/211 (6.2) | 8/160 (5.0) | 5/51 (9.8) | 0.312 | 2.06 [0.5-7.54] | 3/65 (4.6) | 10/146 (6.8) | 0.758 | 1.52 [0.37-8.87] |
| Lymphocytes <1,000 mm3 | 171/210 (81.4) | 126/158 (79.7) | 45/52 (86.5) | 0.311 | 1.63 [0.65-4.69] | 44/65 (67.7) | 127/145 (87.6) | **0.001** | 3.35 [1.54-7.35] |
| Platelets <150,000 mm3 | 46/208 (22.1) | 28/156 (17.9) | 18/52 (34.6) | **0.020** | 2.41 [1.12-5.14] | 11/63 (17.5) | 35/145 (24.1) | 0.364 | 1.50 [0.68-3.55] |
| Hemoglobin <10 g/dl | 13/210 (6.2) | 10/158 (6.3) | 3/52 (5.8) | 1.000 | 0.91 [0.15-3.71] | 8/64 (12.5) | 5/146 (3.4) | **0.024** | 0.25 [0.06-0.91] |
| C-reactive protein >10 mg/liter | 78/210 (37.1) | 49/159 (30.8) | 29/51 (56.9) | **0.001** | 2.94 [1.47-5.97] | 11/63 (17.5) | 67/147 (45.6) | **<0.001** | 3.93 [1.84-9.05] |
| Procalcitonin >0.5 ng/ml | 13/111 (11.7) | 7/79 (8.9) | 6/32 (18.8) | 0.191 | 2.35 [0.59-9.05] | 1/28 (3.6) | 12/83 (14.5) | 0.178 | 4.52 [0.61-202.01] |
| Lactate dehydrogenase >250 U/liter | 115/189 (60.8) | 81/146 (55.5) | 34/43 (79.1) | **0.007** | 3.02 [1.30-7.68] | 26/54 (48.1) | 89/135 (65.9) | **0.032** | 2.08 [1.04-4.16] |
| Aminotransferase aspartate >40 U/liter | 79/191 (41.4) | 55/147 (37.4) | 24/44 (54.5) | 0.055 | 2.00 [0.96-4.21] | 18/56 (32.1) | 61/135 (45.2) | 0.108 | 1.74 [0.87-3.57] |
| Alanine aminotransferase >40 U/liter | 61/188 (32.4) | 41/143 (28.7) | 20/45 (44.4) | 0.067 | 1.98 [0.93-4.19] | 14/53 (26.4) | 47/135 (34.8) | 0.302 | 1.48 [0.70-3.27] |
| Total bilirubin >1.0 mg/dL | 58/170 (34.1) | 41/133 (30.8) | 17/37 (45.9) | 0.116 | 1.90 [0.84-4.27] | 17/48 (35.4) | 41/122 (33.6) | 0.858 | 0.92 [0.44-2.00] |
| Creatine kinase >200 U/liter | 4/23 (17.4) | 2/15 (13.3) | 2/8 (25.0) | 0.589 | 2.09 [0.12-35.61] | 1/7 (14.3) | 3/16 (18.8) | 1.000 | 1.37 [0.09-84.6] |
| Creatinine >1.5mg/dL | 17/210 (8.1) | 10/158 (6.3) | 7/52 (13.5) | 0.139 | 2.29 [0.70-7.12] | 2/63 (3.2) | 15/147 (10.2) | 0.103 | 3.45 [0.76-32.03] |
| D-dimer >500 g/liter | 86/165 (52.1) | 60/128 (46.9) | 26/37 (70.3) | **0.015** | 2.66 [1.15-6.51] | 18/44 (40.9) | 68/121 (56.2) | 0.112 | 1.85 [0.87-3.99] |
| Sodium <134 mg/dL | 33/211 (15.6) | 26/160 (16.2) | 7/51 (13.7) | 0.826 | 0.82 [0.28-2.12] | 6/64 (9.4) | 27/147 (18.4) | 0.148 | 2.17 [0.82-6.78] |
| Potassium <3.5 mg/dL | 41/202 (20.3) | 28/156 (17.9) | 13/46 (28.3) | 0.145 | 1.80 [0.77-4.06] | 16/61 (26.2) | 25/141 (17.7) | 0.185 | 0.61 [0.28-1.34] |

Statistically significant differences in bold (p<0.05).
† 227 (70.5%) patients had a chest X-ray (70.5%). The alterations with a frequency < 5 patients were: pneumothorax (n=2) and pleural effusion (n=1). Chest X-ray results were not available in 5 patients.
‡ 28 (8.7%) patients had a chest CAT scan. Alterations with a frequency of < 5 patients were: pulmonary thromboembolism (n=4), emphysema (n=4), lobar pulmonary infiltrates (n=2), pneumonia (n=2), pneumothorax (n=1), atelectasis (n=1) and pleural effusion (n=1). CAT scan results were not available in five patients.

### Treatment, complications and evolution

Treatment included hydroxychloroquine in 162 (50.3%) patients, azithromycin in 149 (46.3%), lopinavir/ritonavir in 132 (40.7%), glucocorticoids in 34 (10.6%) and tocilizumab in 27 (8.4%), among others (**Table 4**). 49.1% of patients required hospitalization. The mean hospital stay was 9.4 (SD 5.8) days. Phone follow up was made in 277 (86.0%) non-hospitalized and discharged patients, 57 (17.7%) patients were monitored at home. 161 (77.8%) of the 207 patients of working age sought work disability due to COVID-19. The ICU admission rate was 13.0%. The evolution included adult respiratory distress syndrome in 37 (11.5%) patients, severe renal failure in 8 (2.5%), pulmonary thromboembolism in 4 (1.2%) and sepsis in 3 (0.9%) patients. Occupational contact with persons with confirmed or suspected COVID-19 infection was reported by 71 (22.0%) patients, while 51 (15.8%) reported that contact occurred in the family setting. Occupational contact was a protective factor against hospitalization (OR = 0.23; 95%CI = 0.12 to 0.42), ICU admission or death (OR = 0.05; 95%CI = 0.00 to 0.31). The mortality rate to date was 5.6%.

**Table 4.** Predictors of the evolution, complications and treatment in patients hospitalized or with ICU admission/death.

| Variables | Total<br>(n=322) | Death or ICU admission (n=56) |  |  |  | Hospitalization (n=158) |  |  |  |
| --- | --- | --- | --- | --- | --- | --- | --- | --- | --- |
|  |  | No (n=266) | Yes (n=56) | P | OR [95% CI] | No (n=164) | Yes (n=158) | P | OR [95% CI] |
| **Hospital stay— days†** | 9.4 ± 5.8 | 8.9 ± 5.2 | 11.0 ± 7.3 | 0.161 |  | 9.4 ± 6.4 | 9.4 ± 5.6 | 0.990 |  |
| **Complications — no. (%)‡** |  |  |  |  |  |  |  |  |  |
| Any complication | 195 (60.6) | 139 (52.3) | 56 (100.0) | **<0.001** | Inf [12.98-Inf] | 47 (28.7) | 148 (93.7) | **<0.001** | 36.28 [17.28-84.14] |
| Pneumonia | 177 (55.0) | 128 (48.1) | 49 (87.5) | **<0.001** | 7.50 [3.23-20.36] | 44 (26.8) | 133 (84.2) | **<0.001** | 14.35 [8.11-26.19] |
| Adult respiratory distress syndrome | 37 (11.5) | 6 (2.3) | 31 (55.4) | **<0.001** | 52.31 [19.22-168.10] | 10 (6.1) | 27 (17.1) | **0.003** | 3.16 [1.42-7.61] |
| Renal failure | 8 (2.5) | 4 (1.5) | 4 (7.1) | **0.033** | 5.00 [0.90-27.75] | 1 (0.6) | 7 (4.4) | **0.034** | 7.52 [1.00-342.05] |
| Pulmonary thromboembolism | 4 (1.2) | 3 (1.1) | 1 (1.8) | 0.536 | 1.59 [0.03-20.24] | 0 (0) | 4 (2.5) | 0.057 | Inf [0.69-Inf] |
| **Treatments — no. (%)¶** |  |  |  |  |  |  |  |  |  |
| Hydroxychloroquine | 162 (50.3) | 125 (47.0) | 37 (66.1) | **0.012** | 2.19 [1.16-4.26] | 45 (27.4) | 117 (74.1) | **<0.001** | 7.49 [4.48-12.76] |
| Azithromycin | 149 (46.3) | 120 (45.1) | 29 (51.8) | 0.380 | 1.31 [0.70-2.43] | 42 (25.6) | 107 (67.7) | **<0.001** | 6.06 [3.66-10.19] |
| Lopinavir/Ritonavir | 131 (40.7) | 100 (37.6) | 31 (55.4) | **0.017** | 2.05 [1.10-3.86] | 30 (18.3) | 101 (63.9) | **<0.001** | 7.86 [4.61-13.69] |
| Oxygen therapy | 86 (26.7) | 48 (18.0) | 38 (67.9) | **<0.001** | 9.50 [4.83-19.31] | 18 (11.0) | 68 (43.0) | **<0.001** | 6.09 [3.33-11.62] |
| Intravenous antibiotics | 77 (23.9) | 48 (18.0) | 29 (51.8) | **<0.001** | 4.85 [2.52-9.38] | 18 (11.0) | 59 (37.3) | **<0.001** | 4.81 [2.61-9.22] |
| Glucocorticoids | 34 (10.6) | 16 (6.0) | 18 (32.1) | **<0.001** | 7.33 [3.23-16.87] | 5 (3.0) | 29 (18.4) | **<0.001** | 7.11 [2.62-24.20] |
| Tocilizumab | 27 (8.4) | 17 (6.4) | 10 (17.9) | **0.013** | 3.17 [1.22-7.88] | 7 (4.3) | 20 (12.7) | **0.008** | 3.24 [1.27-9.35] |
| Cephalosporins | 22 (6.8) | 18 (6.8) | 4 (7.1) | 1.000 | 1.06 [0.25-3.40] | 1 (0.6) | 21 (13.3) | **<0.001** | 24.82 [3.89-1034.40] |
| Low molecular weight heparin | 19 (5.9) | 12 (4.5) | 7 (12.5) | **0.030** | 3.01 [1.00-8.79] | 1 (0.6) | 18 (11.4) | **<0.001** | 20.82 [3.21-874.82] |
| Remdesivir | 6 (1.9) | 4 (1.5) | 2 (3.6) | 0.280 | 2.42 [0.21-17.36] | 2 (1.2) | 4 (2.5) | 0.441 | 2.10 [0.30-23.52] |
| **Covid-19 infection — no. (%)** |  |  |  |  |  |  |  |  |  |
| Any cohabitant | 51 (15.8) | 39 (14.7) | 12 (21.4) | 0.227 | 1.58 [0.70-3.40] | 25 (15.2) | 26 (16.5) | 0.879 | 1.09 [0.58-2.09] |
| Any work colleague | 71 (22.0) | 70 (26.3) | 1 (1.8) | **<0.001** | 0.05 [0.00-0.31] | 57 (34.8) | 14 (8.9) | **<0.001** | 0.23 [0.12-0.42] |
| Any contact person in other settings | 11 (3.4) | 6 (2.3) | 5 (8.9) | **0.026** | 4.22 [1.00-17.32] | 2 (1.2) | 9 (5.7) | **0.032** | 4.87 [1.00-47.05] |

Statistically significant differences in bold (p<0.05).
† Hospital stay calculated in 122 discharged patients.
‡ Complications in < 5 patients were: sepsis (n=3), multiorgan failure (n=2), electrolyte alterations (n=2), hematologic alterations (n=2) and lung cancer (n=1).
¶ Treatments with a frequency of < 10 patients, except remdesivir, were: amoxicillin (n=6), interferon (n=5), situximab (n=5), darunavir (n=2) and entecavir (n=1).

## DISCUSSION

This study summarizes the clinical, biological and radiological characteristics, evolution and prognostic factors of patients with COVID-19 disease. To date, we are aware of only one published Spanish study [10], which reported on ICU admissions in a region where the epidemic was reported early. Although there have been two systematic reviews and meta-analysis that analyse the clinical characteristics of COVID-19, they are limited to Chinese cohorts or case series [11,12] and a large USA cohort [13] that did not analyse clinical predictors of a poor prognosis.

Clinically, the same main symptoms of cough and fever are reported in all series. However, in Barcelona city, we have observed diarrhoea, anosmia and dysgeusia, which is hardly reported in the Chinese series [7] which, unlike ours comes principally from hospitals: diarrhoea occurred in 23.8% of cases, very similar to the 23% in New York [14] and clearly higher than the 3.8% reported in China. Nearly 20% of patients had anosmia and dysgeusia, similar to the results obtained in French patients [15]. Surprisingly, both were factors of a good prognosis. In contrast, expectoration was found in only 9%, compared with 33.7% in the Chinese series.

Chinese patients had a mean age of 47 years, ten years lower than our series, and 35.7% of our patients were aged ≥ 65 years, compared with 15%, 29% and 31% in China, Germany and the USA respectively, but > 40% in Italy [7,16-18]. Older age and male sex predisposed to a higher mortality rate in our and all large series [7,13]. Dyspnoea, auscultatory alterations and oxygen saturation < 92% were also clinical factors of a poor prognosis. In our patients, comorbidities were three times higher than in the Chinese cohort [7] and were similar to the findings of the New York study [13,14]. The same comorbidities were identified, with hypertension and diabetes being the two most common, while in the USA and Italy, obesity seems to be higher.

Strikingly, 38.2% of our patients were healthcare workers, compared with 3.5% in Wuhan and 5.2% in Germany [7,16]. Although these studies recognized an important degree of underreporting of cases in health workers, the difference remains important. There are at least two possible explanations: first, the lack of personal protective equipment in the initial phase of the epidemic, a constant revindication of health professionals, who felt undersupplied. Secondly, many cases were health professionals from primary healthcare or the reference hospital who reside in the same area where they work.

In all reported series, bilateral pneumonia was the most common radiological finding, was present in more than half the cases [19] and was a factor of a poor prognosis and mortality. In contrast, an interstitial radiological pattern did not confer an increased risk of mortality. The Wuhan study reported a CAT scan use of 88.7%, compared with 8.7% in Barcelona. In contrast, chest X-rays were carried out in 59.1% and 70.5%, respectively: the availability of diagnostic means was higher in China. A recent international consensus states that radiological assessment is not necessary in asymptomatic patients or those with mild disease but is required in patients with moderate or severe disease, regardless of whether a definite diagnosis of COVID-19 has been made [20]. In addition, simple chest radiology [20] is preferable in a resource-constrained environment with difficulties in accessing CAT scans [20]. The possible use of pulmonary ultrasound for the point-of-care diagnosis of COVID-19 pneumonia has not been sufficiently analysed but might be an efficient alternative due to its portability and reliability [21]. In fact, the regional Catalan government has recently acquired 90 ultrasound machines to enable family physicians to make doctors can make point-of-care (home or nursing home) diagnoses of pneumonia [22]. Biologically, lymphopenia and increased CRP, LDH and D-dimer were usually constant and similar in all series and, together with a low platelet count were associated with an increased risk of mortality. A differential variable in our series is a greater number of alterations in liver tests, which was present in 30-40% of patients, data similar to the USA and Italian cohorts, but different from the Chinese cohort, where it was 22% [7]. We also found hypokalaemia in 20.5% of patients, a factor not reported in other studies.

We found a hospitalization rate of 48.7%, compared with 20-31% in the USA and 93.6% in China, and an ICU admission rate of 13%, which was similar to the Chinese (15%), USA (5-11.5%) and German (10%) results. While the protocols of action and admission are similar and depend on the level of clinical involvement, the therapeutic protocols differ between hospitals, cities, and countries. There remain many unknowns in the treatment of COVID-19. The only truth is that we do not have a vaccine, an etiological treatment or a treatment with sufficient scientific evidence to generalize its use. Currently, the systematic review of antiretroviral treatments has not offered conclusive results [23] and despite possibly encouraging in vitro results for hydroxychloroquine, COVID-19 infections are currently intractable [24,25]. The mortality rate in our study was 5.6%, compared with 10.2% in New York (21% in hospitalized patients), 1.4% in China, 3.1% in Germany and 6.8% in Italy. Different information and recording systems, the availability of diagnostic tests, and above all, the organization of national health systems may have contributed to the differences observed.

The study had some limitations due to the observational, retrospective design. However, it is sufficiently representative of the population with confirmed COVID-19 to permit better identification of the factors of a poor prognosis of the disease from a clinical perspective.

Three months after the declaration of the pandemic, there is not a sufficiently reliable, available and generalizable diagnostic test that can analyse the seroprevalence of COVID-19, even in the most industrialized countries. Given this lack, determining the clinical, biological and radiological characteristics of probable cases of COVID-19 infection will be key to the initiation of early treatment and isolation, and for contact tracing, especially in primary healthcare.

## Data Availability

The data that support the findings of this study are available from the corresponding author upon reasonable request.

## ACKNOWLEDGEMENTS

The authors wish to thank David Buss for his editorial assistance.

## ETHICAL APPROVAL

The study was approved by the Ethics Committee of the Hospital Clinic of Barcelona (HCB/2020/0525).

